# Safety-Stock: Predicting the demand for supplies in Brazilian hospitals during the COVID-19 pandemic

**DOI:** 10.1101/2020.05.27.20114330

**Authors:** Oilson Alberto Gonzatto, Diego Carvalho Nascimento, Cibele Maria Russo, Marcos Jardel Henriques, Caio Paziani Tomazella, Maristela Oliveira Santos, Denis Neves, Diego Assad, Rafaela Guerra, Evelyn Keise Bertazo, José Alberto Cuminato, Francisco Louzada Neto

**Affiliations:** Department ofApplied Mathematics and Statistics, Institute of Mathematical and Computer Sciences, University of São Paulo, São Carlos, Brazil; Bionexo, São Paulo, Brazil

**Keywords:** COVID-19 Pandemic, Outbreak, Healthcare Supply Chain, stock-out mitigating risk, easy-to-use free expert system.

## Abstract

**Background:** Many challenges lie ahead when dealing with COVID-19, not only related to the acceleration of the pandemic, but also to the prediction of personal protective equipment consumption to accommodate the explosive demand. Due to this situation of uncertainty, the hospital administration encourages the excess stock of these materials, over-stocking products in some hospitals, and provoke shortage in others. The number of available personal protective equipment is one of the three main factors that limit the number of patients at a hospital, along with the number of available beds and the number of professionals per shift.

**Objective:** In this scenario, a challenge is to build an easy-to-use expert system to predict the demand for personal protective equipment in hospitals during the COVID-19 pandemic, which can be updated in real-time for short term planning.

**Methods:** We propose naive statistical modeling, which combines historical data of the consumption of personal protective equipment by hospitals, current protocols for their uses and epidemiological data related to the disease, to build predictive models for the demand for personal protective equipment in Brazilian hospitals during the pandemic. We then embed our modeling in the free Safety-Stock expert system, which can provide the safety stock levels for a particular hospital.

**Results:** The Safety-Stock system provides the prediction of consumption/demand for personal protective equipment over time, indicating the moment when the hospital reaches maximum consumption, the estimate of how long it will work in this state, and when it will leave it.

**Conclusion:** With our predictions, a hospital may have its needs related to specific personal protective equipment estimated, taking into account its historical stock levels and possible scheduled purchases. The tool allows for the adoption of strategies to control and keep the stock at safety levels to the demand, mitigating the risk of stock-out. As a direct consequence, it also enables the interchange and cooperation between hospitals, aiming to maximize the availability of equipment during the pandemic situation.

## 1. Introduction

As of June 6th, 2020, more than 6.6 million confirmed positive cases of COVID-19 worldwide, with 392,000 global deaths and more than 673,000 confirmed cases in Brazil [1]. Hospital systems around the world have been overwhelmed by the volume of cases, shortages of personal protective equipment (PPE), critical medical supplies, and increasing costs [2]. In this scenario of scarce hospital resources, a challenge is to build an easy-to-use expert system to predict the demand for PPE in hospitals during the COVID-19 pandemic.

Indeed, due in large part to the increase in demand, technological solutions for process management that use historical data from hospital supplies are necessary for the healthcare area. Factors such as price increases and difficulties in purchasing critical supplies without a long term demand prediction require intelligent maintenance of stock levels so that there is no shortage of supplies in hospitals. It is worth noting that the supply chain costs account for the second-largest category of expenses in a hospital, being only lower than labor costs [3].

Aiming to predict the demand for PPE in Brazilian hospitals during the pandemic, we propose naive statistical modeling, which combines historical data on the consumption of PPE, current protocols for their uses, and epidemiological data related to the disease. We then embed our modeling in a free expert system, hereafter Safety-Stock, that presents predictions of consumption/demand for PPEs overtime in real-time. The Safety-Stock indicates the time when the hospital reaches its maximum consumption, estimates how long it will work in this state and when it will leave it. The structure of our modeling is graphically summarized in Figure 1.

**Figure 1:**
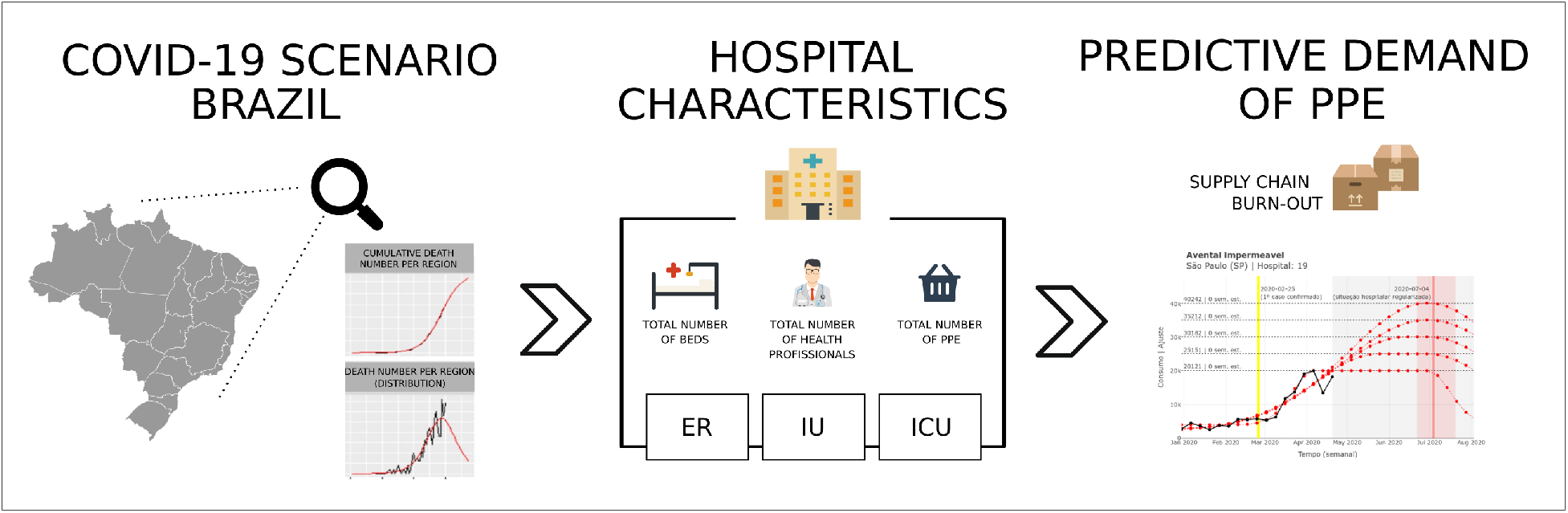
Overview of the structure used in the proposed modeling. The process adopted by the algorithm is to estimate the number of infected patients which will demand some kind of assistance and, combined with the hospital capacities and characteristics, predict the PPE consumption during the COVID-19 pandemic.

With our prediction, a hospital may estimate, based on its stock levels and future purchases, its need related to a specific PPE. This estimate allows for the adoption of strategies to keep stock levels that are adequate to the demand, mitigating the risk of stock-out. As a direct consequence, it also enables the interchange and cooperation between hospitals, aiming to maximize the availability of equipment during the pandemic situation.

The paper is organized as follows: Section 2 presents the materials and methods, including data and statistical modeling. Section 3 describe the Safety-Stock system and the results of its application in several different Brazilian hospitals. Final comments in Section 4 complete the paper.

## 2. Material & Methods

Our modeling uses parametric prediction models to meet the PPE demand of a hospital-based on the estimated epidemic curve and the hospital’s characteristics, such as admission rate, number of emergency department visits, and the total number of available beds. We propose a mathematical/statistical model that expresses the expected relationship of the consumption of a given PPE over time, with the epidemiological characteristics of the region and the internal features of a particular hospital.

Three fundamental pieces of information are taken into account by the modeling process: the recent historical record of hospital consumption of their critical PPEs, the maximum possible level of PPE consumption, the magnitude of the persistence in a maximum consumption regime plateau. Each of these characteristics requires the observation of different sources of information and generates meaningful interpretations for the construction of the model.

For the development of the proposed approach, the following characteristics related to a given hospital are considered:

- List of PPE and other healthcare supplies, such as hand sanitizer, waterproof aprons, sterile gloves, procedure gloves, surgical masks, N95 masks, and caps.
- Availability of these supplies in stock at the hospital, in the form of units in stock.
- Forecast of weekly consumption, which can be calculated in two ways. The first is considering the expected number of hospitalizations of the particular hospital and the consumption of inputs per hospitalization. The second is considering the consumption according to the hospital’s occupancy rate (offer as calculated, as we consider the curve). In addition to consumption, it is necessary to know how these data are calculated, and the hospital must check their calculation.
- The number of available separated or not by the type of case (mild, severe, and critical). The average length of stay for each type of bed is used for the simulation over longer periods. The hospital must provide both the number of beds and their average occupancy rates.
- Value of the hospital safety stock, which represents the minimum stock of each equipment as calculated by the hospital.
- Initial conditions: current hospital occupancy rate and an estimate of the length of stay for critical and non-critical beds.

The curves related to demand estimated by each city of Brazil were constructed using a growth curve model to meet demand from the worst-case situation. We also used information related to the hospital concerning PPE and its infra-structure for occupation. Moreover, we consider the prediction of the demand for hospitalizations and the number of health staff.

The statistical model that parameterizes the gathered data simulates the consumption of resources, and aims at obtaining an indication of the safety stock for each PPE, considering the possibility of having a certain amount. Further details of the statistical modeling can be found in Appendix A.

## 3. Results

The proposed methodology was implemented in Safety-Stock, an easy-to-use expert system that combines different elements informed by the hospital under analysis. It is freely available in https://cemeai.shinyapps.io/ bionexo_covid19/ (in Portuguese). The pieces of information needed for feeding the platform are divided into three dimensions: *consumption and stock information*, which are hospital general information, such as, location (state and city), PPE general classes, weekly consumption, current stock level; *disease behavior in the hospital under study*, which are hospital pandemic dynamic data, such as percentage of hospitalized infected patients, percentage of ICU hospitalized patients, the average length of stay, number of beds and occupancy rate; *forecast of scenarios related to demand*, which are hospital demand prediction data, such as prediction horizon, security percentage, and maximum consumption.

The first step is to estimate the death growth rate related to each region. A growth curve function is adopted to predict the number of deaths in the city where the hospital lies. Examples for these curve functions include the Three-Parameter Logistic [4, chap. 6] and the Gompertz Function [5], for instance. Figure 2a illustrates this estimate by showing the cumulative number of deaths in five major Brazilian cities: Belo Horizonte (MG), Recife (PE), Curitiba (PR), Porto Alegre (RS) and São Paulo (SP).

**Figure 2:**
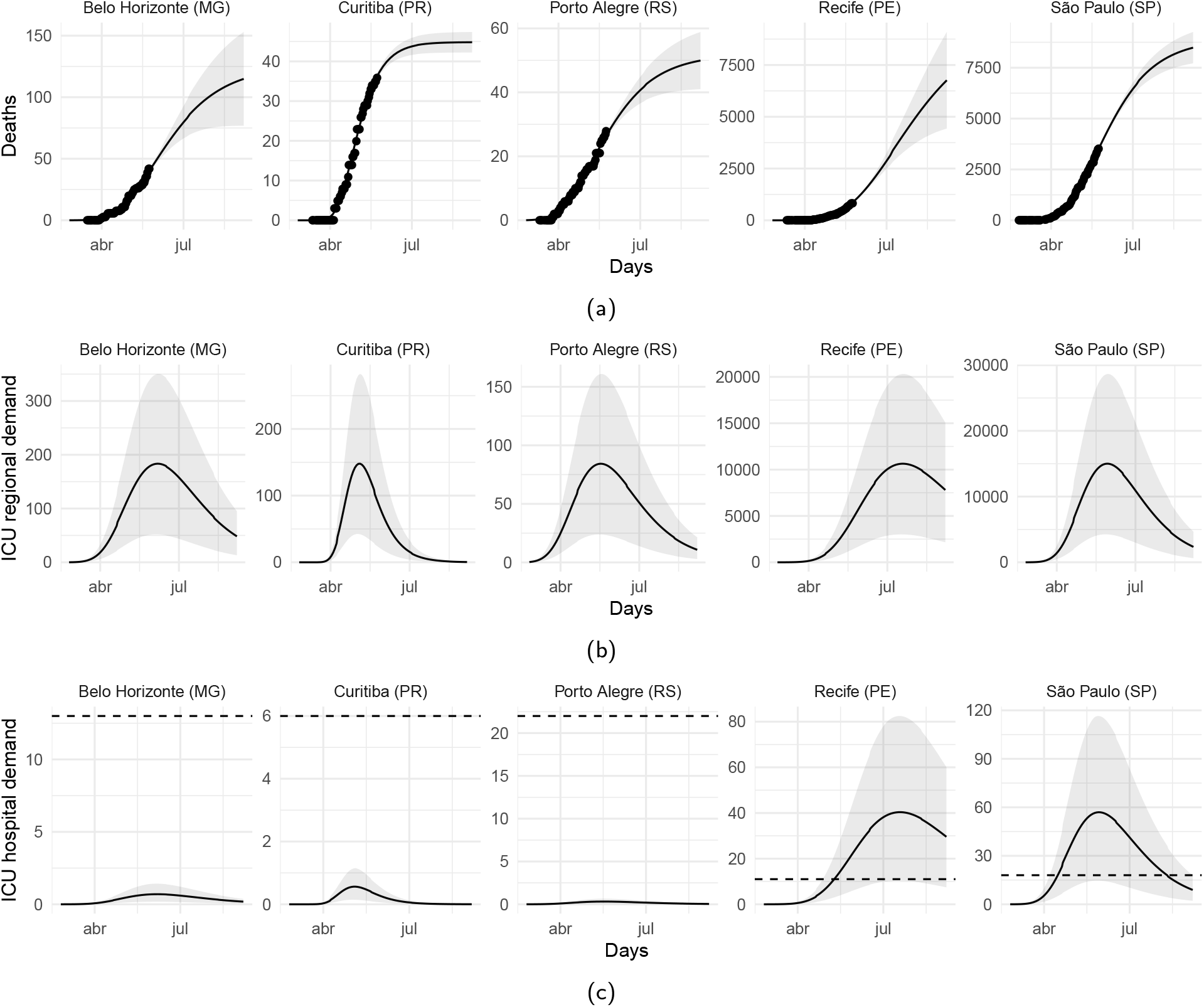
Pandemic dynamic estimation for five Brazilian cities. The selected cities are Belo Horizonte (MG), Recife (PE), Curitiba (PR), Porto Alegre (RS) and São Paulo (SP). The top panel (a) represents the cumulative death rate per region, the middle panel (b) expresses the expected dynamics of the disease and the bottom panel (c) represents the market share fraction expected to be attended by the analyzed hospital from that city.

Then, based on the process described in Appendix C, we obtain the behavior for regional demand for each city (Figure 2b), which expresses a particular behavior concerning the dynamics of the disease. The fraction of such curve that fits a specific hospital depends on the market share associated with it, and the cut-line that represents its current capacity (in terms of the number of free ICU beds) considers the total number of beds and the respective occupancy rates (Figure 2c). With this step, we determine the cut-line that gives us an indication of when the hospital situation begins to regularize.

Subsequently, a simulated hospital structure, which is focused exclusively on the COVID-19 pandemic, is considered. This simulated hospital’s composition was an essential element for estimating the threshold that relates to the maximum consumption of PPE: the number of available beds, the size of medical staff, and the PPE consumption protocols. With this step, we defined a technically standardized level for maximum consumption, on which we established some variation in its surroundings to contemplate possible changes in the protocol due to the pandemic situation. A detailed description of the simulated hospital structure is found in Appendix B.

The next step consists in adjusting the conditioned model to the information obtained in the previous steps. Thus, we established a forecast for the weekly PPE consumption of a hospital under analysis through models considering some possible scenarios. Figure 3 presents the estimate in which the pandemic presents a potential risk to the hospital situation as a result. The generated scenarios consider the historical consumption of a PPE in particular, the theoretical premises for the use of PPE based on the magnitude of the hospital, and the effects of the pandemic observed in its region. The gray margin exposes the forecast horizon.

**Figure 3:**
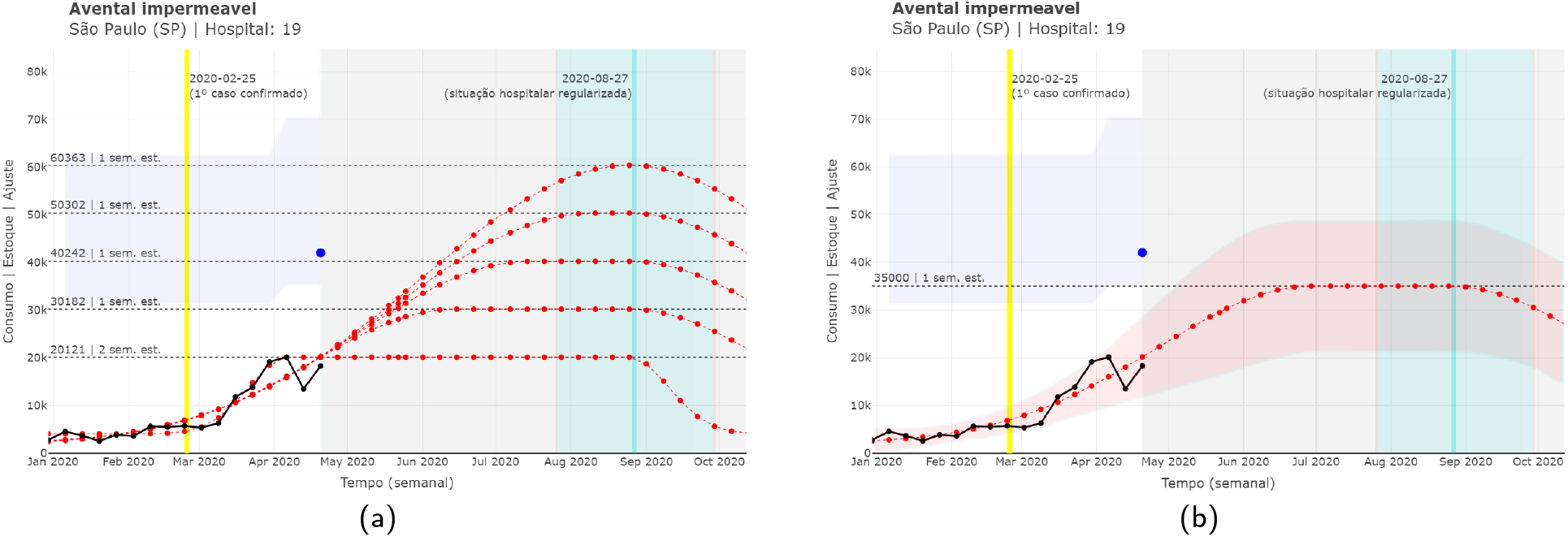
Dynamic estimation towards a PPE, considering the simulated hospital. The black line and dots represent the consumption history available in the spreadsheet. The dashed lines (horizontal) in black represent possible consumption limits. Left-hand panel (a) displays the different PPE level demands based on the hospital capacities, meanwhile right-hand panel (b) displays the chosen option according, e.g. to the supply chain manager. There figures were taken directly from the online platform, which is available in Portuguese.

In Figure 3, each consumption limit line also indicates the security of the available stock in terms of the number of weeks that the current stock would last if there were no new entries. The red dashed lines represent upper limits for consumption, considering the growth trend of previous records and different total consumption levels, the transparent red margins represent the 2.5% and 97.5% limits for the predicted curve. The blue dot denotes the current stock position. The transparent blue margins represent the minimum and maximum stock limits. The yellow line showed the date when the first case of COVID-19 was identified in its municipality. The vertical line in light-blue represents a cut-line from which we believe that the hospital situation will return to normal. The transparent margin around it expresses the uncertainty involved in this expectation.

This value for maximum consumption will be given as input to the model (submitted in the Maximum Consumption field), and it will be recalculated, making the specific analysis to hospital reality (3b). The red line is the upper limit for consumption, where growth occurs with the intensity outlined by the historical record and considering its consumption protocol.

This growth ceases from the moment the hospital situation begins to regularize (in this example, this occurs by late August 2020). The construction of this cut-line takes into account the situation of the hospital while facing the pandemic.

The board of the hospital under analysis, in the face of the pandemic, may use the cut-line as an aid in decision making, estimated by the considered approximations and assumptions, supporting the hospital’s regularization situation by the following steps:

1. Based on the death count due to COVID-19. We obtain an estimate for this behavior. Then, we add other sources of uncertainty, such as under-reporting factors.
2. Conditioning the real death curve, we consider the estimated relationship between the death and infected curve, added by a randomness factor.
3. Subsequently, via the infected curve, estimates of the fraction of hospitalized and, of those hospitalized, those who need ICU admission.
4. From the curve of intensive care unit inpatients, we considered the average number of days in the intensive care unit, and, given this consideration and the randomness involved, we took an approximation, even if gross, of the curve of recoveries.
5. The difference between the intensive care unit inpatients curve and the recoveries curve leads us to a hospital demand curve for intensive care unit beds.

The fraction of the curve applied to the hospital under analysis is considered an estimate for market-share.

The first panel (left) in Figure 4 shows an estimate of the death curve of Brazilian municipalities (extracted from the website brasil.io), later this growth curve helped in the corrected estimation bypassing the underreporting of COVID- 19, see more details in the appendices. The central panel is related to the region’s demand curve, and finally, the right panel adds the daily demand of the hospital (being a fraction of the region answered by market share).

**Figure 4:**
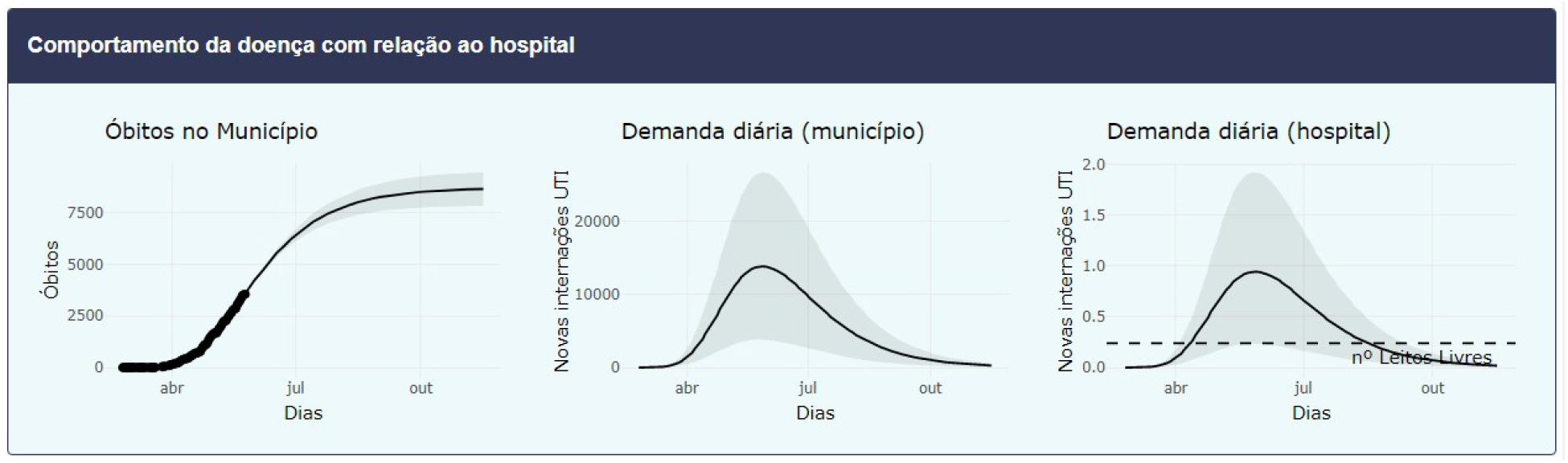
Pandemic dynamic per region based on the analyzed hospital. Left-hand panel plots the death growth, center panel related to the region's demand curve, and right-hand panel display the daily estimation demand of the hospital. Online platform print, which is available in Portuguese.

The total sum will represent the Forecast Horizon can be increased by a percentage defined in the % security field. The fragmentation of this forecast according to the percentage of consumption of each item individually, in the last 30 days.

As an auxiliary tool, the expected demand for each group of commonly used items in the pandemic will be broken down into individual products (with different references). This table can be exported and aims to consider the different scenarios that relate to the maximum consumption.

Perhaps an essential point of the Safety-Stock is the accumulated record of hospitals that need to be helped, and those that can help others. This initiative can be materialized with an indicator that tells us if the hospital can help, or needs help, after analyzing the situation of lack or over-stock for each general class of PPE or the need to purchase more supplies.

To learn about the Safety-Stock flow of information, interested readers can refer to Appendix D, where we illustrate the flow of information that runs internally in our expert system, according to Figure 5.

**Figure 5:**
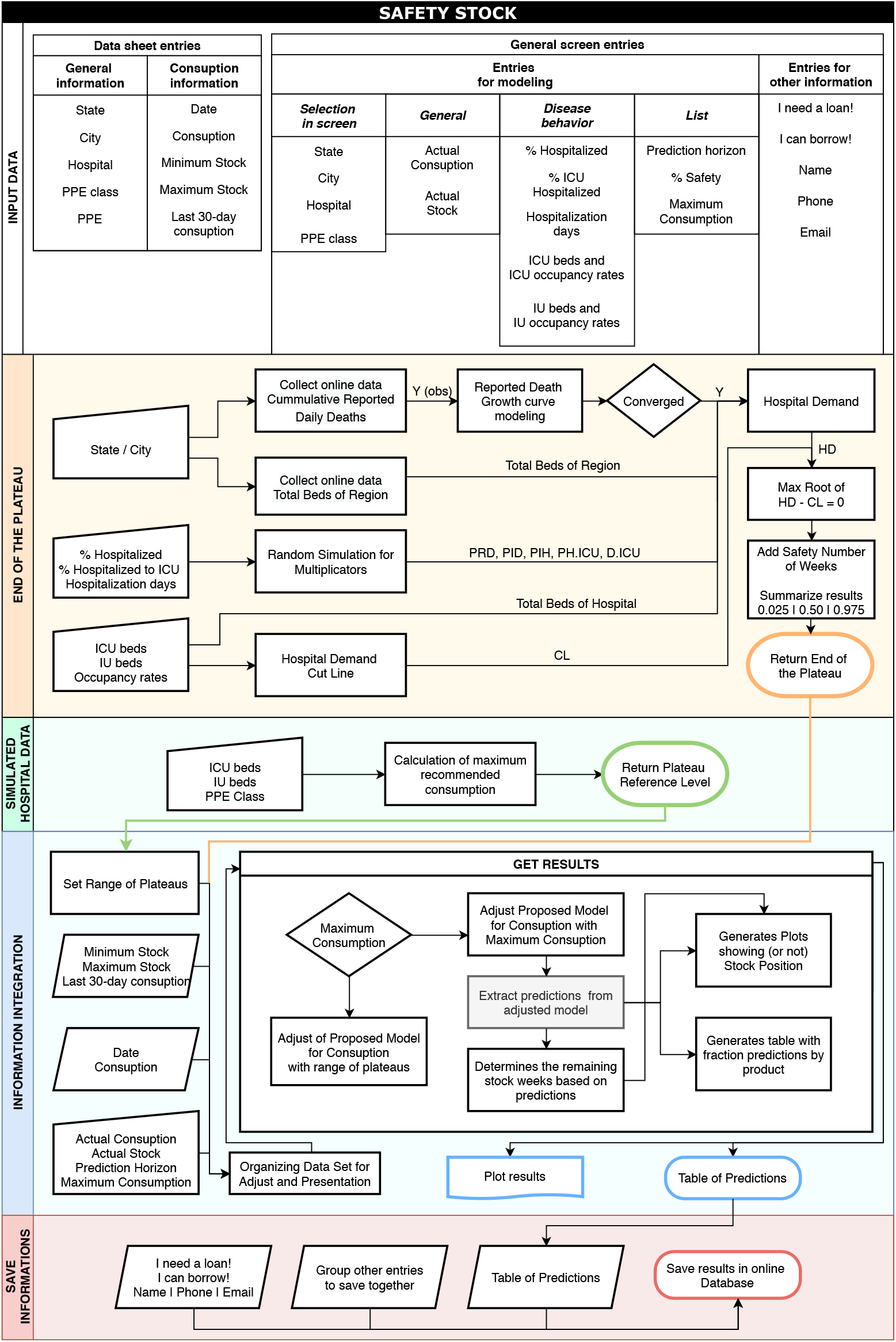
Safety-Stock information flow.

## 4. Final Comments

Prediction modeling was developed by combining historical hospital data and the disease growth curve. The idea is to provide a safety stock, avoiding a possible lack of PPE during the COVID-19 pandemic. We developed the Safety-Stock system, which provides to the hospital managers a prediction for the consumption of several PPEs, taking into account the expected number of patients that arrive in the emergency room with COVID-19 symptoms, as well as the expected percentage of those that need intensive care. As a result, the safety stock of PPEs can be estimated. Consequently, it may be rearranged among geographically close hospitals, preventing attendance restrictions, and avoiding unnecessary expenses.

The proposed approach is naive in different ways. For instance, we chose to assume a random structure based on a symmetric probability distribution for the errors, both in the adjust of the consumption curve (*C_t_*) and the death curve *(Y_t_)*. The choice of such distributions may not be the most appropriate one, though entirely consistent results were found. In other words, the raw data directly accessed in the statistical estimation processes correspond to the recent history of the consumption of any PPE and the death record in a particular city. All other information carries subjectivities and uncertainties that we cannot quantify in light of the analyzed data. Even though the theoretical support of the studies, from where such information was extracted, allowed us to understand the observed average behaviors as premises in our modeling. Finally, the considered growth models are quite simple and describe the disease’s mean behavior over time.

All of these points can be refined and addressed in future studies. For instance, assign a possibly asymmetric probability distribution for the errors, consider joint statistical modeling the various information, with data directly accessed by us, and consider other growth models and then use statistical selection among different models.

On the other hand, naivety also has some attractive advantages. Scalability is one of them since complex models commonly require computationally intensive methods, many of them need a more substantial amount of information to express good results and the computational processing time is considerably longer. In this sense, our naive approach allows the use and diffusion of the proposed methodology on a broader spectrum of possibilities, such as the use of our expert system with updating in real-time. Another interesting point is that researchers from other areas can clearly understand the methodology, and its results can be effectively internalized. Besides, the integrated set of small independent solutions, such as the proposal presented here, can serve as a basis for more in-depth investigations by expressing insights that are difficult to perceive in individual analyzes.

Moreover, results obtained with the Safety-Stock were exposed to some PPE managers of some Brazilian hospitals. The degree of agreement with the reality of their practical activities encourages us the continuity of the development,maintenance, and dissemination of our research and free expert system.

## Data Availability

There is no statement regarding the availability of any data.

## Appendix A – Statistical Methodology

Consumption/demand curve. In principle, we consider a random variable *C_t_* denoting the *consumption of a given PPE over time t*, normally distributed with mean *μ_t_* and variance *σ*^2^.

The adopted statistical model aims to parameterize the growth behavior of the demand for a product, followed by a (possibly) period of constant high demand, then accompanied by a decline in demand. Thus, *μ_t_* is assumed as a five parameters function given by

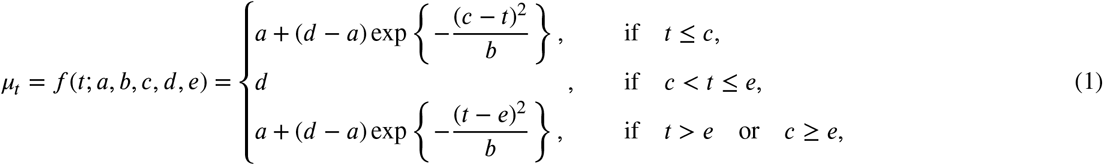

where *a* denotes the magnitude of basic consumption, here assumed to be constant, *b* denotes the intensity of growth/ decrease in consumption over time, *c* denotes the point in *t* that the maximum consumption is reached, *d* denotes the magnitude of maximum consumption, *e* denotes the point in *t* that consumption begins to decrease.

According to the variation of parameters *a, b, c, d* and *e*, some possible behaviors for the mean of the proposed model can be observed in Figure 6.

**Figure 6:**
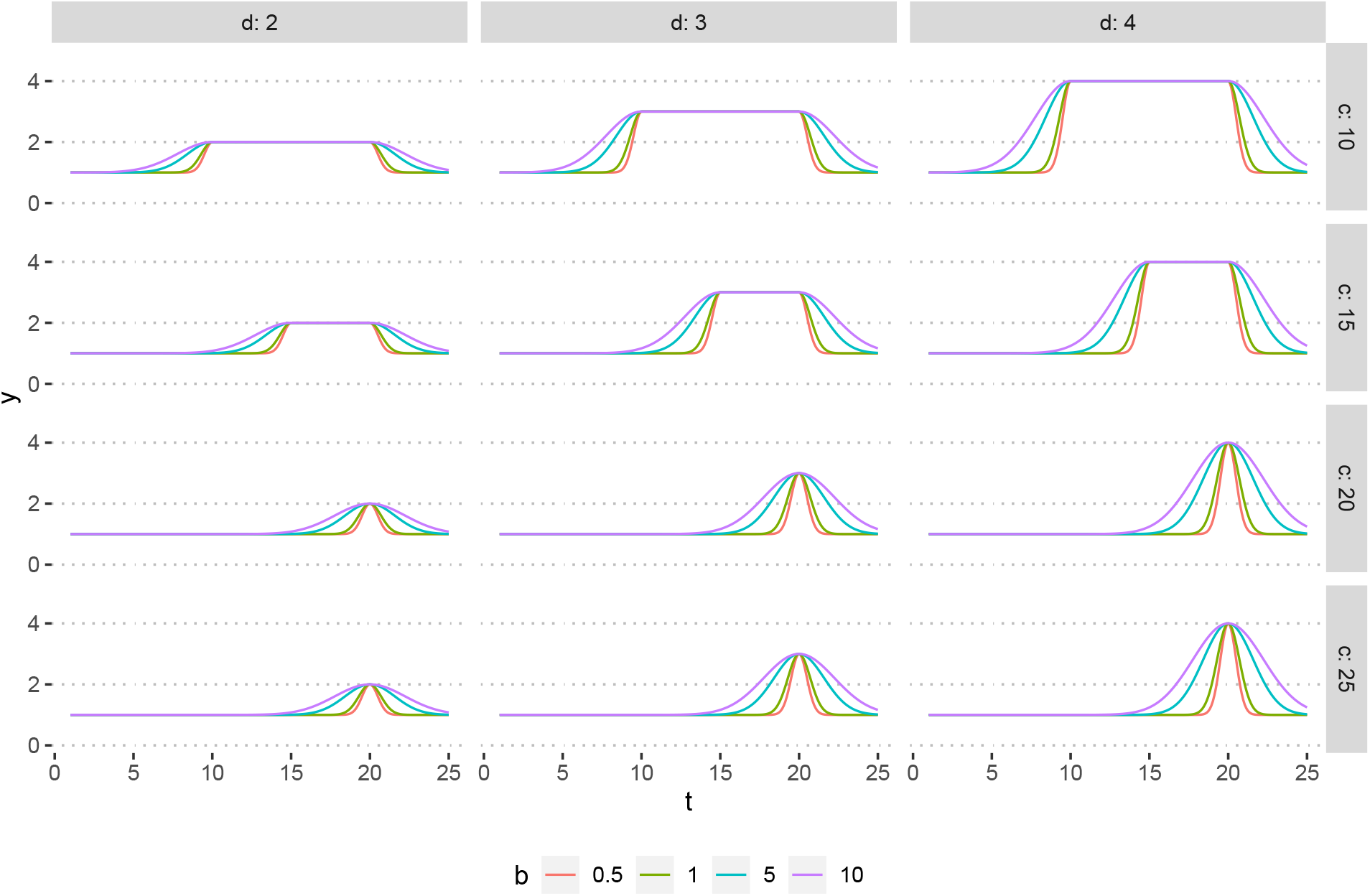
Some possibly behaviors of *mu_t_* for fixed values of *a* = 1, *e* = 20 and *b* = 0.5,1.0,1.5,10, *c* = 10,15,20,25 and *d* = 2, 3,4.

The described proposal that relates the model *C_t_* with the consumption curve with the regional epidemiological characteristics and interior features of a particular hospital takes into account three fundamental aspects. The recenthistorical record of hospital consumption of a given PPE, The maximum consumption level of the PPE, How long the hospital stays on the maximum consumption level. Each of these features requires the observation of distinct information sources and generate meaningful interpretations for the model building, as follows.

1. The recent historical record of hospital consumption of a given PPE provides information of the baseline demand before the COVID-19 pandemic (represented by the model parameter *a*) and evidence of changing in the consumption regime (represented by the model parameter *b*);
2. The maximum level of PPE consumption considers the maximum capacity and the totality of the hospital staff dedicated to COVID-19 patients care (this maximum level is represented by the parameter *d* of the model and, as it is a fixed and particular characteristic of each hospital and PPE, it is set as a known parameter). This information is determined based on the procedure described in Appendix B;
3. The time spent on the maximum consumption regime (whose endpoint in *t* is represented by the *e* parameter of the model) is determined by the use of hospital information and the region around it. It is estimated indirectly and is subsequently used in the model for the consumption curve, *C_t_*, as a fixed parameter (just like the parameter *d*). The determination of the *e* parameter, defined as known in the model for *C_t_*), is done as described in Appendix C.

The combined use of information obtained from these three perspectives makes the theoretical model adopted to relate the epidemiological characteristics of the region and the demand features of the hospital’s supply chain under study.

In this sense, as the parameters, *d* and *e* are determined indirectly (see Appendices B and C) therefore considered known in the curve expressed by *μ_t_*, the parameters *a, b* and *c* must be estimated by some estimation process.

Estimation Process. We define the random structure, based on a set of observations from recent consumption history 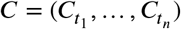, 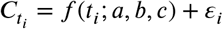, where 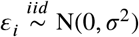, for *i =* 1,…, *n*.

The parameters *θ = (a, b, c)* e *σ*^2^ are estimated by considering the log-likelihood function, given by

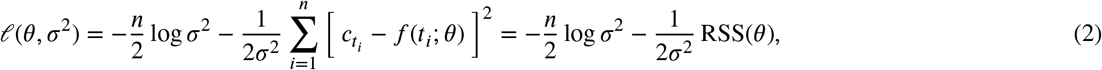

where RSS(*θ*) denotes the residual sum of squares.

If we use non-informative or sufficiently vague a priori information for the parameters, estimates for *θ* and *σ*^2^ can be obtained by maximizing (2), which occurs with the minimization of RSS(*θ*) (*θ* is independent of *σ*^2^). In addition, *∂ℓ/∂σ*^2^ = 0 has a solution given by 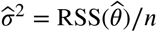, while 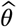 is the least squares estimator of *θ* [6]. The standard error of the estimates can be obtained based on the Fisher Information matrix and the prediction intervals are determined using the Delta Method [7].

## Appendix B - Simulated Hospital Data

In order to evaluate the proposed model, a simulated hospital environment was considered, based on ANVISA, the Brazilian Health Regulatory Agency [8]. This simulation serves as a general example for the hospitals where the model will be applied, and its characteristics (staff and material consumption) were defined using data provided by experts.

The hospital allocates its patients into three categories: Inpatient Units (IU), Intensive Care Units (ICU), and Emergency Room (ER). IU patients are in a non-critical state, while UTI patients are in a critical state, demanding more human resources and materials. The number of hospital beds is divided between both units, and IU beds can be turned into ICU if needed. ER patients are on hold to be transferred to either IU or ICU. Therefore they are not considered to be occupying hospital beds.

The allocation of doctors, nurses, and physiotherapists is shown in Table 1. The number of IU and ICU needed staff is given based on the number of occupied beds, except for doctors for IU patients, which is given by the number of total hospital beds, regardless of their occupation. Column IU/RRT shows the staff allocated to IUs as Rapid Response Team (RRT). In these columns, the numbers represent the staff needed for 12-hour shifts, while numbers on the ER column represent the daily staff needed for a unit with an average of 10.000 treatments per month, which also work on 12-hour shifts.

**Table 1.** Staff allocation in the simulated hospital.

| Staff | IU | IU/RRT | ICU | ER |
| --- | --- | --- | --- | --- |
| Doctors | 1 / 10 total beds | 1 / 100 beds | 1 / 10 beds | 20 |
| Nurses | 1 / 6 beds | 1 / 100 beds | 1 / 8 beds | 10 |
| Physiotherapists | 1 / 20 beds | 1 / 100 beds | 1 / 10 beds | - |

IU e ICU: values for a 12-hour shift; ER: daily values for an unit with 10.000 monthly treatments.

Table 2 shows the consumption of critical material per professional during a 12-hour shift. These values estimate what is used taken from a series of premises and observations on real hospitals.

**Table 2.** Material consumption in the simulated hospital.

| Material | Doctor | Nurse | Physiotherapist |
| --- | --- | --- | --- |
| Surgical Mask (unit) | 6 | 6 | 6 |
| Waterproof Apron (unit) | 1 | 2 | 2 |
| Hospital Cap (unit) | 1 | 1 | 1 |
| Procedure Gloves (pair) | 5 | 10 | 10 |
| Sterile Gloves (pair) | 2 | - | 20 |

Average values per professional in a 12-hour shift.

The consumption of other essential materials needs to be estimated by other means. The use of Hand Sanitizer varies from hospital to hospital since it can be replaced by regular soap. In this case, it is assumed the daily use of 20mL for each IU and ICU patient and 5mL for each ER suspected patient (which is estimated as 50%). Doctors, nurses, and physiotherapists use N95 Masks, at a rate of 1 each 14-day period or ten shifts, thus depending on the staff rotation rather than hospital occupancy.

Lastly, as a premise, the obtained values are increased by 40% to consider material usage from cleaning and technical staff, visitors, and incoming patients, as well as hospital waste.

## Appendix C - Calculating the end of the plateau

For the determination of a cut-line, from which the number of beds available in the hospital becomes again higher than the daily demand of patients in ICU (represented by the parameter *e* in the model (1)), we used the information of the number of notified deaths accumulated over time (here denoted by the random variable *Y_t_)* and other hospital information as described below.

The growth curve of *Y_t_* can be estimated by a nonlinear growth curve model, such as the Logistic model [4], Gompertz, Richards [5], Von Bertalanffly [9], among others. The selection between the most appropriate model for each region can be made based on some statistical criteria such as AIC, AICc, BIC [10]. The estimation process is analogous to what is described in Appendix A.

From the estimated curve for *Y_t_*, we consider a brief simulation study to approach the *hospital demand* curve. We incorporate other sources of uncertainty, such as underreporting factors and lethality rates, among others. The characterization of such sources of uncertainty has theoretical support in studies already published, which support the average behavior defined here as a known premise. This approach may represent a current study limitation since we do not contemplate the joint analysis of all the factors involved, which can be incorporated for future studies, though such information has already been obtained and validated by other sources.

Thus, the steps performed are the following:

- The *reported deaths* curve *(Y_t_)* is a fraction of the *actual deaths 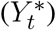*, which means that

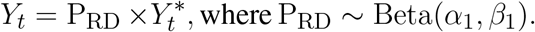 The probability distribution of P_RD_ (proportion of reported deaths) has been defined so that we consider the parameters *α*_1_ and *β*_1_ to be known, such that 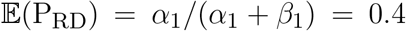 and 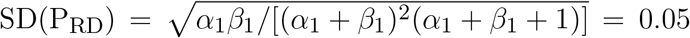. Such assumptions take into account researchers such as [11, 12] which indicate that the number of *real deaths* is around 2.6 times the number of *reported deaths*.
- We understand that the *real deaths* curve 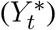 is proportionally related to the *infected* curve *(I_t_)*, as follows,

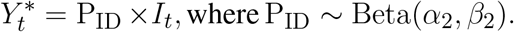 The probability distribution of P_iD_ (proportion of infected-to-death) was defined considering known *α*_2_ and *β*_2_, such that 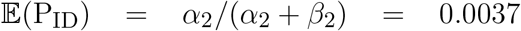 and 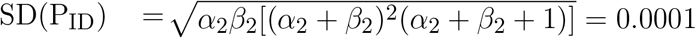. The distribution parameter of the random variable P_ID_ was set based on the results of [13, 14] which pointed out that the actual lethality rate of the disease is around 0.37%;
- The *hospitalized (H_t_)* and *hospitalized in ICU (H_ICU,t_)* curves are assumed to correspond to fractions of the curve of *infected*, as follows

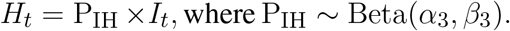

and

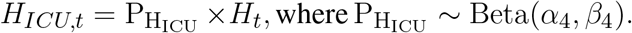 Similarly, the probability distribution of P_IH_ (proportion of infected-to-hospital) and 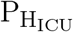 (proportion of hospital- ized-to-ICU) are defined as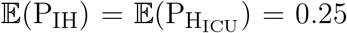 and 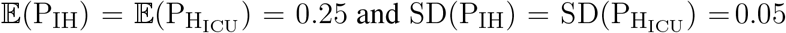. This is justified since [15, 14] shows that approximately 25% of the infected need some hospital care, and 25% of those need a ICU.
- The *recovered* curve *(R_t_)*, from ICU situation, was approximated by the translate of the difference between *hospitalized in ICU* curve *(H_ICU,t_)* and the *real deaths* curve 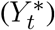, as follows [16]

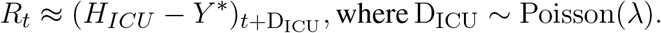 The probability distribution set to D_icu_ (days in ICU) takes into account that 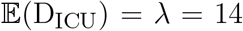, since [17] indicated that the clinical recovery for patients in ICUs is approximately two weeks.
- In addition, we understand that the difference between the *hospitalized in ICU* curve *(H_ICU,t_)* and the *recovered* curve (*R_t_*) of the ICU situation approximates the *regional demand* curve (*RD_t_*) for ICU beds. Therefore

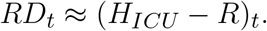
- Finally, we assume that the *hospital demand* curve *(HD_t_)* is a fraction of the *regional demand* curve (*RD_t_*), weighted by market share (MS) associated with a particular hospital. Therefore

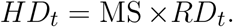 The multiplier MS was not considered as a random variable. Its value is determined by the number of beds in the hospital and the number of beds available in the region close to the hospital, whose information is collected directly from a Brazilian health database called DataSUS.

Once we approximate the *hospital demand* curve (*HD_t_*), we have established a cut-line for the hospital’s service capacity. This procedure is made by considering the number of IU and ICU beds (named as IU_B_ and ICU_B_, respectively) and their occupancy rates (named as OR_IU_ and OR_ICU_, respectively). Since around 20% of IU beds can be upgraded to act like an ICU bed, the cut-line is given by

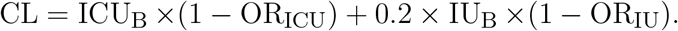

Thus making *HD_t_* = CL, we identify the cutoff point, *t*^(1)^, which indicates that the ICU service capacity has been exceeded (if applicable) and the point at which it has returned to normal is *t*^(2)^.

The point *t*^(2)^ and all the uncertainty associated with it, via the variability of P_RD_, P_ID_, P_IH_, 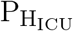, and D_ICU_, result in a range of possible time points, where the hospital situation is expected to return to normal. The average value of these estimates is set as the cut-line represented by the parameter *e* of model (1) (assumed known in this stage of the modeling).

## Appendix D - Safety Stock toll information Flow

In this appendix, we illustrate the flow of information that runs internally on the Safety-Stock system (Figure 5). All input data is listed in the INPUT DATA panel. The remaining processes are divided into four large blocks: 1) The END OF THE PLATEAU panel outlines the cut line estimation process from which we understand that the hospital situation begins to regularize. 2) The SIMULATED HOSPITAL DATA panel represents the stage of determining the maximum consumption level used as a reference in the later stage. 3) The INFORMATION INTEGRATION panel shows the flow of joining the information obtained previously, for the construction and presentation of the solution. 4) Finally, the SAVE INFORMATION panel condenses the records that must be saved in the online database.

In the INPUT DATA panel, a categorization of the entries can be seen. The first one deals with the datasheet loaded by the user of the Safety-Stock and corresponds to the identification information (of the region, hospital, and PPE class), the historical consumption record, and the stock requirements. All other information is provided via the Safety-Stock screen. Some of them deal with empirical aspects about the disease’s behavior in the region of the hospital (or the hospital itself), such as the average number of days that a patient remains in the ICU, or other epidemic information that is particularized in specific health centers. All the above information is defined by default (regarding recent medical literature) at the beginning of the user’s section. However, it can (and should) be modified to meet the specific practical reality of that hospital.

Other necessary entries deal with some small configurations about the forecast stage of the model, such as the forecast horizon and the safety percentage added.

The second frame, END OF THE PLATEAU, link the flow of information used to determine the cut line represented by the parameter *e* of the model (1). Considering the region selected by the user, a module of the Safety-Stock searches online for the death record in that region, updated daily. Besides, there is a query in the dataSUS database about the number of beds available in the given location. Another module adjusts a growth curve to the data collected for the cumulative death number. Some transformations will be made in order to obtain the hospital demand curve. The transformations are made with the aid of some multipliers (which weigh the death curve, P_IH_, 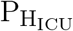, P_ID_, among others), and a factor that translates the curve over time (D_ICU_). The multipliers are obtained by a module via random simulation, considering the exposed in Appendix 4. Having access to the hospital’s demand curve in another block of code, we determined some possible dates for the plateau’s end, considering the cut line obtained based on the entries associated with the number of beds and respective occupations.

In the third panel, SIMULATED HOSPITAL DATA, the reference for maximum consumption level is obtained by a module that makes use of information on the number of total beds, and the product class. The dimension of this plateau takes into account all aspects listed in Appendix 4.

The fourth table expresses the joint use of the information obtained in Tables 1 to 3. The *maximum consumption* stipulated by the user indicates which stage of the analysis he is. If this entry is NULL, then the consumption curve adjustment model considers a range of plateaus based on the reference plateau (obtained in third 3). On the other hand, if the user has already defined the *maximum consumption*, the model fits a single curve, associated with that consumption, at the end of the analysis, this will be the curve used to make and save the predictions. Other modules are in charge of generating the prediction table and the graph that outlines the adjusted model compared to the observed data and other relevant information.

The last panel summarizes the information that will be exported to the company’s database responsible for an intermediate between hospitals that self-declared as potential helpers (for the loan or exchange of PPE) and those who declared that they need help.

## Notes

* The research was carried out using the computational resources of the Center for Mathematical Sciences Applied to Industry (CeMEAI), funded by FAPESP (grant number 2013/07375-0). José Alberto Cuminato, Francisco Louzada and Maristela O. Santos are supported by the Brazilian agency CNPq (grant number 302954/2015-4, 301976/2017-1 and 306305/2018-6, respectively).

### Competing Interest Statement

The authors have declared no competing interest.

### Funding Statement

The research was carried out using the computational resources of the Center for Mathematical Sciences Applied to Industry (CeMEAI), funded by FAPESP (grant number 2013/07375-0). Jose Alberto Cuminato and Francisco Louzada are supported by the Brazilian agency CNPq (grant number 302954/2015-4 and 301976/2017-1, respectively).

### Author Declarations

There is no IRB/oversight body.

